# How does DeepSeek-R1 perform on USMLE?

**DOI:** 10.1101/2025.02.06.25321749

**Authors:** Lisle Faray de Paiva, Gijs Luijten, Behrus Puladi, Jan Egger

## Abstract

DeepSeek, a Chinese artificial intelligence company, released its first free chatbot app based on its DeepSeek-R1 model. DeepSeek provides its models, algorithms, and training details to ensure transparency and reproducibility. Their new model is trained with reinforcement learning, allowing it to learn through interactions and feedback rather than relying solely on supervised learning. Reports showcase that DeepSeek’s model shows competitive performances against established large language models (LLMs) such as Anthropic’s Claude and OpenAI’s GPT-4o on established benchmarks in language understanding, mathematics (AIME 2024) and programming (Codeforces) while trained at a fraction of the costs. Additionally, running inference shows significantly lower costs, leading to DeepSeek surpassing ChatGPT as the most downloaded free app on the American iOS App Store. This development contributed to a nearly 17% drop in Nvidia’s share price, resulting in the most significant one-day loss in U.S. history, amounting to nearly $600 billion. The open-source models also bring a significant shift in the healthcare system, allowing cost-efficient medical LLMs to be deployed within hospital networks. To understand its performance in the healthcare sector, we analyse the new DeepSeek-R1 model on the United States Medical Licensing Examination (USMLE) and compare it to ChatGPT.

## Introduction

Large language models (LLMs) have emerged as a transformative force in artificial intelligence (AI), significantly impacting various domains by enhancing natural language understanding and generation [1]. However, LLMs are often commercial products created by large tech companies such as OpenAI or Google, which means they tend to operate as closed systems. This contrasts with the philosophy of “open source” initiatives, such as those in the medical field, which aim to share knowledge and resources freely with the public. There is a growing concern that access to these powerful tools may be limited to only those who can afford them, including wealthy individuals and researchers, thereby creating a divide in availability, reproducibility and transparency [2].

DeepSeek-R1, a newly developed AI model, represents a significant step forward in natural language understanding and domain-specific reasoning [3, 4]. Unlike proprietary models like ChatGPT, DeepSeek-R1 offers comparable performance while being fully open-source and reporting comparative performances while requiring a fraction of the resources for development. Users can download, deploy, and run it locally, eliminating reliance on cloud-based services and expanding accessibility to a broader audience [3].

Given AI’s critical role in medical education and professional certification, researchers have increasingly explored its potential to assist in medical learning and assessment [5, 6, 7]. With recent advancements in LLMs, AI systems have demonstrated remarkable capabilities in answering complex medical questions, simulating clinical reasoning, and even achieving near-expert performance in medical decision-making [8]. Models such as GPT-4 [9] and BERT [10] are revolutionizing clinical support, diagnosis, treatment, and medical research by processing vast amounts of medical data efficiently [11, 12, 13]. While prior studies have examined the performance of models such as GPT-4 (OpenAI, 2023) [14, 15, 16] and Med-PaLM 2 [17] on medical benchmarks, the effectiveness of DeepSeek-R1 in this context remains unexplored [18].

This study aims to evaluate DeepSeek-R1’s performance on the United States Medical Licensing Examination (USMLE), a standardized assessment used to assess medical students’ and graduates’ knowledge and clinical reasoning [19], by analyzing its accuracy, reasoning capabilities, and potential limitations. We seek to assess its viability as a tool for medical education and clinical decision support. Additionally, we explore the model’s strengths and weaknesses in handling different question types, including clinical vignettes, foundational sciences, and patient management scenarios. Our findings contribute to the ongoing discourse on the role of AI in medical training, with implications for both AI development [20] and specific models and applications, like AI reasoning in pediatric clinical decision support [21].

## Methodology

The evaluation leverages the June 2022 USMLE multiple-choice question (MCQ) dataset from Kung et al. [5], which previously assessed ChatGPT-3’s performance on the exam. This dataset includes questions categorized by USMLE Step (Step 1, Step 2 CK, Step 3), reflecting their distinct focus areas:

- Step 1: Foundational science recall.
- Step 2 CK: Clinical decision-making.
- Step 3: Patient management and synthesis.

Questions were divided into batches of 30 per step. Reference answers from the original study were retained as ground truth.

### Model Evaluation Protocol

For model evaluation, DeepSeek-R1 was prompted with each batch of 30 questions using a zero-shot instruction structure. As reported in the original paper [5], pre-existing outputs from ChatGPT-3 served as the baseline for comparison.

Two complementary metrics were assessed: ROUGE scores [22, 23] and BERTScore [24]. ROUGE-2 measured the exact bigram overlap between model outputs and reference answers, with a 1.0 score indicating verbatim correctness. ROUGE-L evaluated broader textual alignment using the longest common subsequence, with a threshold of ≥0.8 for near-exact matches. BERTScore F1 quantified semantic similarity through contextual embeddings, with scores ≥0.8 reflecting clinically meaningful alignment beyond lexical overlap.

A contamination check was conducted to address potential data contamination. A stratified random sample of 10% of reference answers was selected, and DeepSeek-R1 was prompted to generate corresponding USMLE-style questions. Lexical (word-for-word) and structural comparisons between generated and original exam questions revealed no matches, confirming the novelty of the dataset in DeepSeek-R1’s training corpus.

Statistical comparisons between DeepSeek-R1 and ChatGPT-3 focused on step-wise frequency counts of ROUGE-2 = 1.0 and ROUGE-L/BERTScore ≥0.8, alongside distributional analysis of BERTScore F1 spreads visualized through violin plots. Step-wise partitioning mirrored the USMLE’s hierarchical design, enabling granular analysis of model performance across exam demands.

## Results

The ROUGE and BERTScore metrics analysis reveals differences in DeepSeek and GPT’s performance on USMLE multiple-choice questions. Figure 1 illustrates contrasts in answer accuracy and alignment between the two models across USMLE steps. To contextualize these metrics, Table 1 presents a representative Step 3 question with model outputs and reference answer.

**Table 1:** Comparative example of model responses to a Step 3 question.

|  |  |
| --- | --- |
| Reference Answer | (A) "I am concerned that you are abusing pain medicine. I would like for you to consider a substance use disorder treatment program." |
| DeepSeek Output | (A) "I am concerned that you are abusing pain medicine. I would like for you to consider a substance use disorder treatment program."<br>(RougeL = 1.0 / Bertscore = 1.0) |
| ChatGPT Output | (A) "I am concerned that you are abusing pain medicine. I would like for you to consider a substance use disorder treatment program." It is important to approach the conversation with empathy and concern for the patient's well-being. The best way to start the conversation would be to express concern about the patient's potential substance abuse and to suggest seeking treatment for a substance use disorder. Options B, C, and D are not appropriate ways to approach the conversation, as they may be perceived as confrontational or accusatory, which could lead to the patient becoming defensive or uncooperative. (RougeL = 0.07/ Bertscore = 0.46) [5] |

**Figure 1:**
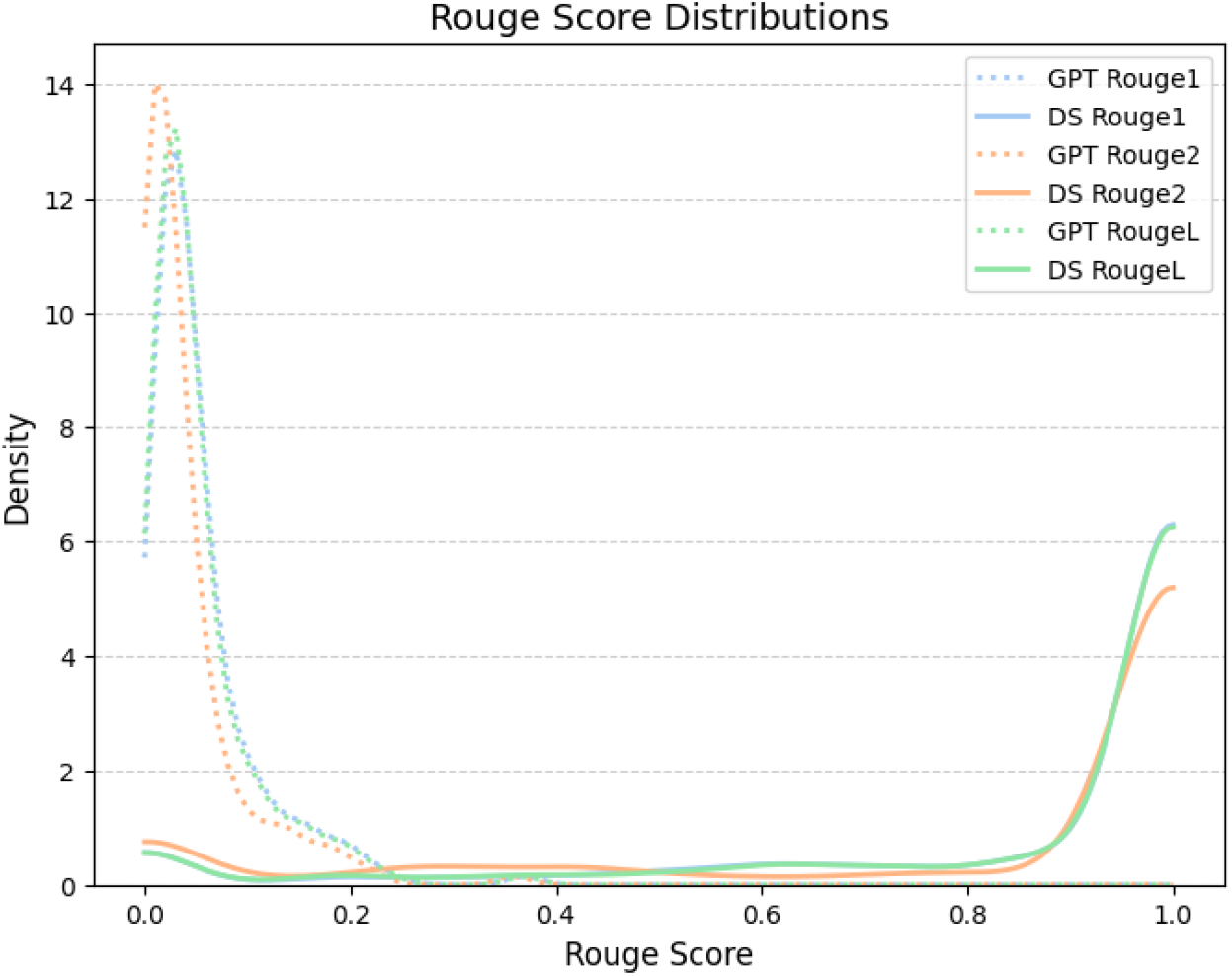
This figure presents the distribution of ROUGE-1, ROUGE-2, and ROUGE-L scores for GPT and DeepSeek (DS) models. The distributions highlight differences in score densities, with both models showing peaks at low and high ROUGE scores. Dashed lines represent GPT scores, while solid lines correspond to DeepSeek scores.

### ROUGE Score Analysis: Model Performance

Figure 1 shows the ROUGE-1, ROUGE-2 and ROUGE-L score distributions across all answers. DeepSeek achieved ROUGE-2 1.0 (indicating exact matches) in 73 out of 94 instances in Step 1, 83 out of 109 in Step 2 CK, and 50 out of 122 in Step 3. GPT did not achieve a ROUGE-2 score of 1.0 in any step. In broader textual alignment (ROUGE-L ≥ 0.8), DeepSeek attained 82 instances in Step 1, 88 in Step 2 CK, and 59 in Step 3. GPT did not reach the ROUGE-L ≥ 0.8 threshold in any step.

### BERTScore Analysis: Semantic Alignment with Reference Text

BERTScore distributions (Figure 2) show DeepSeek achieved semantic similarity scores (F1 ≥ 0.8) in 87 instances for Step 1, 98 for Step 2 CK, and 72 for Step 3. GPT did not surpass the BERTScore F1 threshold of 0.8 in any step.

**Figure 2:**
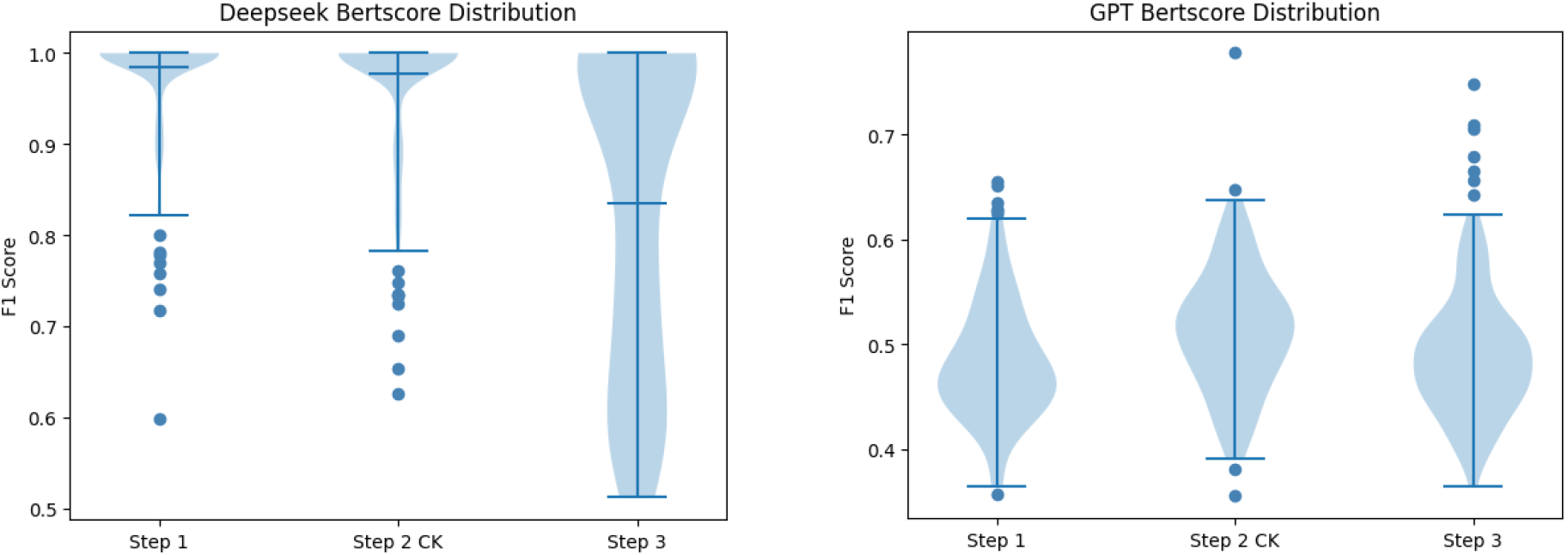
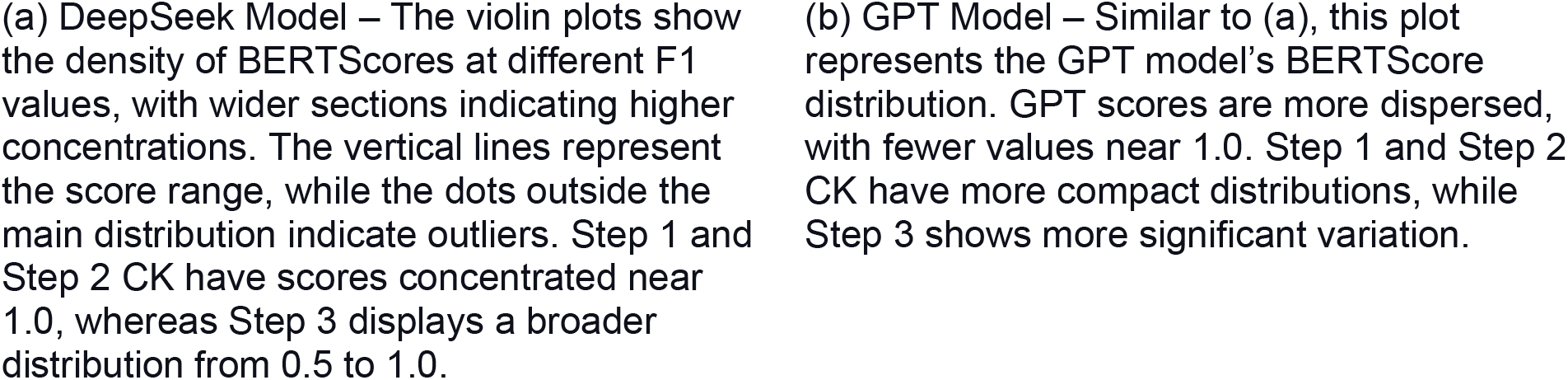
This figure presents violin plots comparing the F1 BERTScore distributions for DeepSeek (DS) and GPT models across three evaluation steps. The violin plots provide insight into score distributions, highlighting concentration differences and the presence of outliers across steps.

### Step-Wise Performance Trends

Step-wise performance trends indicate variability across the USMLE steps. In Step 1, DeepSeek achieved ROUGE-2 exact matches in 73 out of 94 instances. Performance remained strong in Step 2 CK with 83 exact matches out of 109 but decreased in Step 3 to 50 out of 122. GPT exhibited broader BERTScore distributions across all steps compared to DeepSeek.

## Discussion

This study provides insights into the feasibility of large language models (LLMs) in high-stakes medical assessments, revealing stark contrasts between extractive (DeepSeek) and abstractive (GPT) strategies in the USMLE’s precision-dependent format. The DeepSeek-R1 report reinforces this distinction, highlighting the model’s extractive precision in structured reasoning tasks, such as its superior performance in MATH-500 (97.3% Pass@1 - *proportion of correct answers generated on the first attempt*). This makes DeepSeek particularly well-suited for knowledge recall and exact answer retrieval, which aligns with the fact-based demands of Step 1 of the USMLE.

The evaluation reveals stark contrasts between DeepSeek and GPT in answer accuracy and alignment, as shown in Figure 1. DeepSeek demonstrates a strong tendency toward extractive precision, evidenced by its ROUGE-2 scores of 1.0 in 73/94 instances in Step 1, 83/109 in Step 2, and 50/122 in Step 3. This suggests that DeepSeek frequently selects verbatim correct answers. In contrast, GPT failed to achieve a single ROUGE-2 score of 1.0 across all steps, implying it never produced an exact match.

The disparity persists in broader textual alignment (ROUGE-L ≥ 0.8), where DeepSeek excels with 82 instances in Step 1, 88 in Step 2, and 59 in Step 3, indicating its outputs closely mirror reference texts. GPT, however, never reached this threshold, reinforcing its propensity for abstractive summarization with greater paraphrasing and divergence from reference answers. These results highlight DeepSeek’s extractive, reference-aligned approach compared to GPT’s more variable, abstractive strategy.

The BERTScore distributions shown in Figure 2 reinforce DeepSeek’s superior semantic alignment, with 87 instances in Step 1, 98 in Step 2, and 72 in Step 3, achieving a BERTScore F1 ≥ 0.8. This indicates that even when DeepSeek does not produce verbatim matches (as seen in ROUGE-2), its outputs retain high semantic similarity to reference answers. GPT, however, fails to reach the 0.8 BERTScore threshold in any step, underscoring its weaker grasp of contextually or structurally aligned content.

Regarding Step 1 Recall-Driven Context, DeepSeek’s extractive strategy in foundational science questions rewards exact terminology matches. GPT’s abstractive approach fails to meet this rigid standard. On Step 2CK’s decision-making focus, both models perform relatively better. However, DeepSeek’s balance of exact matches (ROUGE-2) and semantic coherence (BERTScore) aligns with clinical reasoning tasks requiring structured logic. Onto Step 3’s Patient Management Complexity, DeepSeek’s exact matches (50/122) decline and increased BERTScore variability suggest clinical ambiguity in this step may demand hybrid strategies (extractive + abstractive), however its outputs remain semantically closer to reference answers than GPT’s. In contrast, GPT’s persistently low scores and wide spreads shown in Figure 2 highlight its inability to consistently approximate correct answers, even semantically.

The hierarchical design of the USMLE, which progresses from foundational recall (Step 1) to clinical synthesis (Step 3), provides a useful framework for interpreting model behaviors. DeepSeek’s dominance in Step 1 (73/94 exact ROUGE-2 matches) highlights its extractive strength in verbatim concept retrieval. However, this advantage diminishes in Step 3, where clinical ambiguity demands adaptive reasoning. Here, its exact matches drop to 50/122, and BERTScore variability increases, suggesting that even extractive models struggle with open-ended patient management tasks. This aligns with DeepSeek-R1’s broader evaluation results [3], where it excels in structured problem-solving (e.g., Codeforces 96.3 percentile) but encounters difficulties in tasks requiring flexible, contextual reasoning. Conversely, GPT consistently fails to achieve ROUGE-L ≥ 0.8 or BERTScore ≥ 0.8, indicating a systemic limitation in medical answer fidelity due to its abstractive preference for paraphrasing over precision.

These findings have clear implications for AI-assisted medical education. DeepSeek’s extractive alignment with reference answers makes it a potent tool for targeted review, particularly in reinforcing foundational concepts in Step 1. For example, medical students preparing for Step 1 could use DeepSeek-powered tools to quickly retrieve precise definitions, biochemical pathways, or pharmacological mechanisms, reducing the cognitive load of memorization-heavy subjects like biochemistry, microbiology, and pathology. In question-based learning (QBL) settings, DeepSeek’s ability to provide exact answer matches could enable automated feedback systems that flag incorrect responses and provide reference-aligned explanations, helping students build a strong factual foundation.

However, its performance in Step 3, though superior to GPT, highlights a key limitation: Clinical decision-making often requires contextual flexibility that pure extractive methods cannot fully capture. In complex patient cases requiring synthesis of multiple clinical factors—such as choosing the best management strategy for a patient with overlapping chronic conditions— DeepSeek’s rigid extractive nature may fail to consider nuances such as patient preferences, evolving symptoms, or atypical presentations. Meanwhile, GPT’s fluency comes at the cost of factual consistency, raising concerns about semantically plausible but clinically incorrect answers in high-stakes scenarios. For instance, an abstractive model might generate a reasonable-sounding but medically inappropriate treatment plan, making it unreliable for decision-support in clinical training.

Finally, the absence of lexical overlap between generated and original USMLE questions supports the conclusion that DeepSeek’s performance reflects genuine reasoning rather than memorization. However, its ability to achieve BERTScore ≥ 0.8 in 72 Step 3 cases despite fewer exact matches suggests that a hybrid architecture—combining extractive precision with controlled abstraction—could better address the complexities of Step 3.

While our analysis provides insights into textual alignment between model outputs and reference answers using ROUGE and BERTscore metrics, these automated metrics primarily assess lexical and semantic similarity rather than clinical accuracy. For a comprehensive evaluation of medical knowledge, particularly in Step 3’s complex patient management scenarios, expert validation by physicians remains critical. As Kung et al. [5] demonstrated, human expert review can identify clinically plausible but factually incorrect reasoning that automated metrics might overlook. Future studies should incorporate physician-led assessments to evaluate diagnostic soundness and the real-world applicability of LLM-generated answers. This dual approach, combining computational metrics with clinical expertise, would better assess the readiness of open-source models like DeepSeek-R1 for deployment in medical education and decision support systems.

### Conclusion

This study provides a comprehensive evaluation of DeepSeek-R1’s performance on the USMLE, highlighting its strengths in extractive precision and structured reasoning compared to the abstractive approach of GPT models. Our findings indicate that DeepSeek-R1 excels in fact-based recall and clinical knowledge retrieval, particularly in Step 1 and Step 2 CK, where its exact-match performance significantly surpasses GPT’s. This suggests its strong potential as a medical education tool, offering accurate reference-aligned responses for targeted review, question-based learning (QBL), and automated feedback systems.

However, DeepSeek’s performance declines in Step 3, where contextual flexibility and adaptive reasoning are essential for managing complex clinical cases. While its outputs remain semantically closer to reference answers than GPT, its extractive approach struggles with the nuances of patient management strategies. Conversely, GPT’s tendency toward paraphrasing and abstraction, though more flexible, comes at the cost of factual consistency, limiting its reliability in high-stakes medical decision-making. These results highlight a critical gap in current LLM architectures— neither pure extractive nor fully abstractive models alone are sufficient for handling the full range of medical reasoning tasks.

The absence of lexical overlap between DeepSeek’s generated responses and original USMLE questions further validates that its performance is driven by genuine reasoning rather than memorization. Nonetheless, our findings suggest that developing a hybrid model, combining DeepSeek’s extractive precision with GPT-like contextual abstraction could enhance clinical decision support systems, adaptive learning environments, and AI-powered tutoring platforms, bridging the gap between structured medical knowledge and real-world clinical reasoning.

Future research should explore how reinforcement learning techniques can optimize DeepSeek-R1’s performance in complex medical scenarios and how hybrid AI models can integrate extractive accuracy with contextual adaptability. Additionally, more models should be evaluated to provide a complete picture of the current state of LLMs in the medical domain.

Finally, while our computational metrics provide objective performance benchmarks, subsequent work should integrate physician evaluation to assess clinical validity, particularly for Step 3’s nuanced patient management challenges.

### Disclaimer

OpenAI’s ChatGPT and DeepSeek’s R1 chat generated hints using customized prompts for some parts of the paper.

## Data Availability

All data produced in the present work are contained in the manuscript

